# Assessing Excess Mortality Patterns in Argentina over the COVID-19 Pandemic (2020-2021): A Comprehensive National and Subnational Analysis

**DOI:** 10.1101/2024.05.31.24308276

**Authors:** Velen Pennini, Adrian Santoro, Santiago Esteban, Camila Volij, Adolfo Rubistein

**Affiliations:** Centro de Implementación e Innovación en Políticas de Salud (CIIPS). Instituto de Efectividad Clínica y Sanitaria (IECS)/Institute for Clinical Effectiveness and Health Policy, Buenos Aires, Argentina

## Abstract

The COVID-19 pandemic has dramatically impacted global health metrics, with the World Health Organization (WHO) reporting over 732 million cases and 6.7 million deaths by the end of 2021. Additionally, approximately 14.8 million excess deaths were estimated globally through 2022, significantly surpassing reported COVID-19 deaths. In Argentina, recorded pandemic-related fatalities reached nearly 160,000 from March 2020 to December 2022, underlining the necessity for a detailed examination of excess mortality across national and subnational levels.

This study aims to describe excess mortality in Argentina in 2020 and 2021 and its subnational geographic areas, and to identify geographic and temporal disparities across sub-regionsusing publicly available monthly mortality and climate data from Argentina, spanning 2015 to 2021. Excess mortality was assessed using Generalized Additive Models (GAM) to account for long-term and annual trends, monthly climatic variations, and epidemiological reports of Influenza-like Illness (ILI). Data across various geographic regions was analyzed to identify temporal and spatial disparities in mortality.

Our analyses revealed significant regional disparities in mortality, identifying a total of 133,612 excess deaths across Argentina during the study period, with notable peaks coinciding with COVID-19 waves. These insights not only contribute to our understanding of the pandemic’s broader effects but also emphasize the critical need for enhanced public health responses informed by mortality data analyses. The development of an open-source, interactive platform further supports this initiative, enabling detailed exploration and informed decision-making to better manage future public health crises.

## Introduction

The global epidemiological impact of the COVID-19 pandemic has been profound, with the World Health Organization (WHO) reporting over 732 million confirmed cases and 6.7 million deaths as of December 31, 2022^1^. Additionally, the WHO estimates indicate that there were approximately 14.8 million excess deaths worldwide through the duration of the pandemic by the end of 2022, a figure 2.7 times greater than the reported COVID-19 deaths during the same period^2^. More recent studies describe that out of 131 million global deaths from all causes combined in 2020 and 2021, approximately 15.9 million were attributed to COVID-19, either directly or indirectly due to associated social, economic, or behavioral changes during the pandemic^3^. In Argentina, the pandemic led to around 10 million confirmed cases^1^ and nearly 160,000 deaths from March 2020 to December 2022, with significant annual variances observed: more than 53,000 in 2020, almost 85,000 in 2021, and almost 24.000 in 2022^4^.

The most substantial impact on the death toll was noted during 2020-2021. Latin America was particularly hard-hit, emerging as one of the regions most affected during this timeframe and several countries in the region presented the highest number of cases and deaths in the world^5,6^. Notably, countries like Peru and Mexico experienced enormous excess mortality rates during 2020, highlighting the severe regional disparities in the pandemic’s impact^7^.

In Argentina, the pre-pandemic general mortality rate remained relatively constant between 809.8 and 757.0 per 100,000 from 2015 to 2019, experiencing dramatic increases to 829.0 and 953.5 per 100,000 in 2020 and 2021, respectively (26.0% increase). However, the assessment of excess mortality has been primarily focused on the year 2020, with limited comprehensive analysis extending into 2021^8–10^.

The anomalous and sudden nature of the pandemic brought significance to the concept of "excess deaths" defined as the number of deaths that occurred in a period exceeding what is expected as "normal" based on historical records. The concept of "excess mortality" is operationalized as a percentage of the total expected deaths under normal conditions and is used as a comparable indicator across different populations, which is useful for quantifying the impact of a pandemic (or other dramatic situations such as climate disasters, wars or catastrophes) on mortality^11^. Unlike other classic mortality indicators, such as crude, adjusted, or specific mortality rates, estimating excess mortality requires the availability of mortality information for all causes for the period of analysis in a given population and a historical period for the same population.

Considering the impact of a pandemic, this indicator is crucial in examining the aftermath, as it considers various factors that can affect fatalities, such as government-driven public health and social policies to manage the crisis, shifts in social behaviors, and preparedness and response of healthcare systems.

While there is consensus that excess deaths observed above what is expected (in absolute values or percentages) represent the appropriate indicator to capture this phenomenon, there is no consensus in the literature on how to determine the threshold of what is expected under "normal" circumstances, and consequently, how to calculate the indicator. Two of the most commonly used alternatives are models based on percentiles of the historical distribution of deaths ^12 8^ and models based on more complex or multivariate methods that can incorporate other variables, such as climate or seasonality ^13–20^.

This study aims to describe excess mortality in Argentina in 2020 and 2021 and its subnational geographic areas, and to identify geographic and temporal disparities across sub-regions. Beyond providing detailed analyses and insights, the research aims to extend its utility by developing an open-source, interactive, customizable, and user-friendly platform for estimating jurisdiction-specific excess mortality. This tool is designed to facilitate the exploration and understanding of mortality data by researchers, policymakers, and the general public, enabling informed decision-making and targeted interventions to address public health challenges.

## Materials and Methods

This is a descriptive and analytic study. Monthly mortality statistical data from 2015 to 2021 were utilized to construct curves representing the monthly number of deaths for all causes at both national and subnational levels. The datasets used were publicly released by the Ministry of Health of Argentina (MoH) and were accessible through its Open Data portal^21^. The available information for excess mortality analysis was temporally disaggregated by month. The geographic areas defined in the dataset encompassed complete jurisdictions in 11 cases (the provinces of Buenos Aires, Chaco, Córdoba, Corrientes, Entre Ríos, Formosa, Mendoza, Misiones, Santa Fe, Tucumán, and the Autonomous City of Buenos Aires). Given the lower event frequency and in order to prevent the indirect identification of deceased individuals, the MoH aggregated the remaining jurisdictions into five regions based on geographic contiguity: Jujuy + Salta (NOA 1), Catamarca + Santiago del Estero (NOA 2), Chubut + Santa Cruz + Tierra del Fuego (Southern Patagonia), La Pampa + Neuquén + Río Negro (Northern Patagonia), and San Juan + San Luis + La Rioja (Rest of Cuyo) (Fig 1).

**Fig 1.**
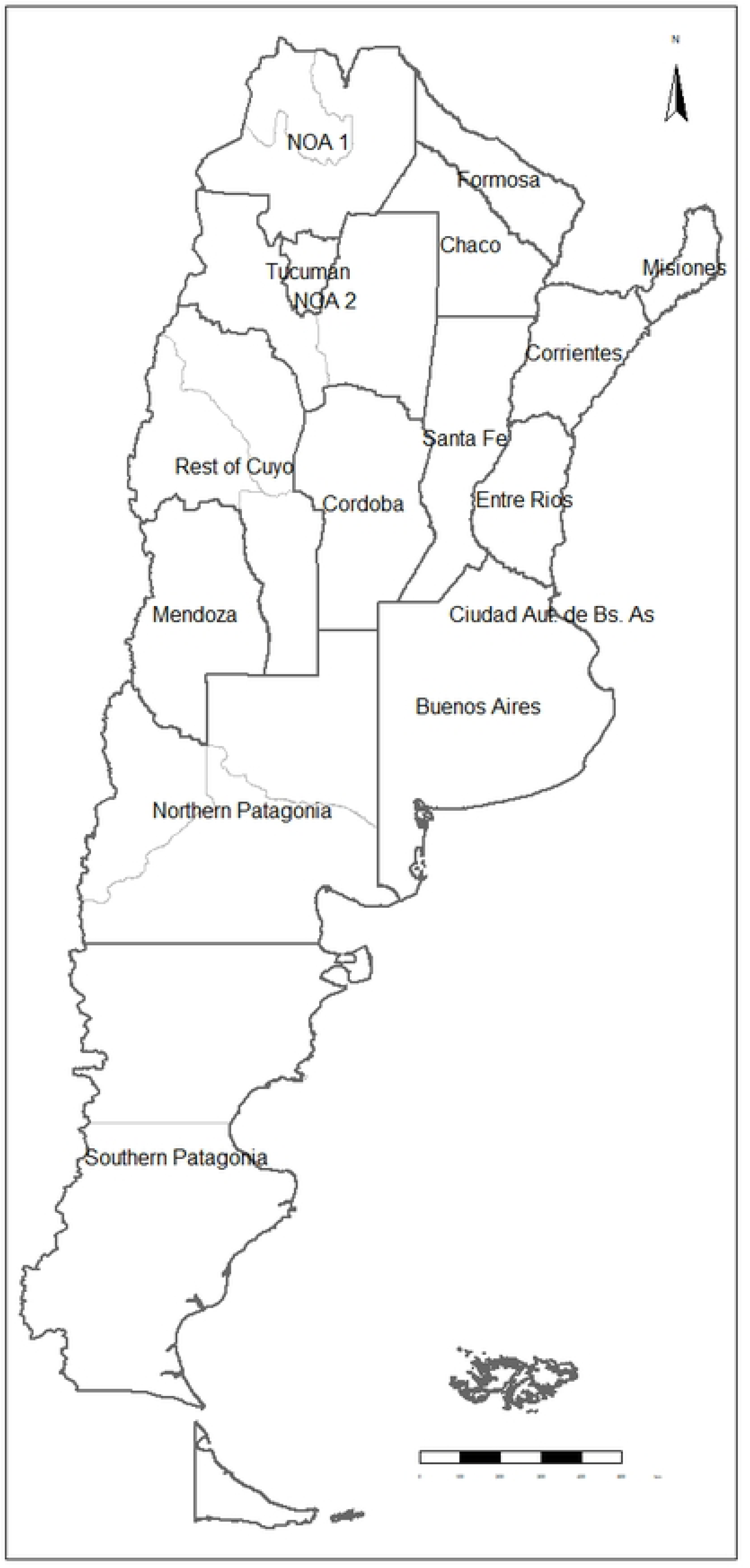
Geographic Division for Excess Mortality Analysis in Argentina.

Additionally, monthly data for both minimum and maximum temperatures^22,23^ recorded by meteorological stations across various geographical regions for the period spanning from 2015 to 2021 were incorporated. Data was obtained from the National Meteorological Service of Argentina^24^. Weekly notifications of Influenza-like Illness (ILI) cases ^25,26^, sourced from epidemiological bulletins periodically published by the Ministry of Health^27^ and the Open Data portal^28^ for years 2015–2021, were also used. Studies cited, employing similar models have demonstrated improved goodness of fit, reinforcing the robustness of our methodological approach.

A generalized additive model (GAM) was identified as the best choice for threshold estimation after evaluating the root mean square error (RMSE) between various models (1) simple linear models, 2) simple linear model with regularization, 3) Poisson regression model, 4) negative binomial generalised linear model, 5) GAM model, and 6) median and percentiles methodology^8^) comparing predicted versus observed mortality within trainning data via cross-validation for the time series over the period 2015-2019. This process was carried out using the R^29^ package caret and the function “trainControl”, with the method specified as "timeslice" ^30^.

During model development and selection, long-term (systematic changes observed throughout the entire study period, independent of seasonal variations) and annual trends, monthly average minimum and maximum temperatures, and reports of ILI cases from 2015-2019 were taken into account. Based on the RMSE obtained through cross-validation, the GAM was chosen as the best estimator to predict the expected number of deaths in 2020-2021 (test set) and to estimate excess deaths (the difference between observed and expected deaths) on a monthly and annual basis, both at the national and subnational levels.

As illustrated in Fig 2, the data was segmented into training, validation, and test sets. The training and validation sets were split sequentially in chronological order, using the last year of each sequence as the validation set. RMSE was used as the metric for selecting the best performing model. The test set (2020-2021) was used for the final performance evaluation.

**Fig 2.**
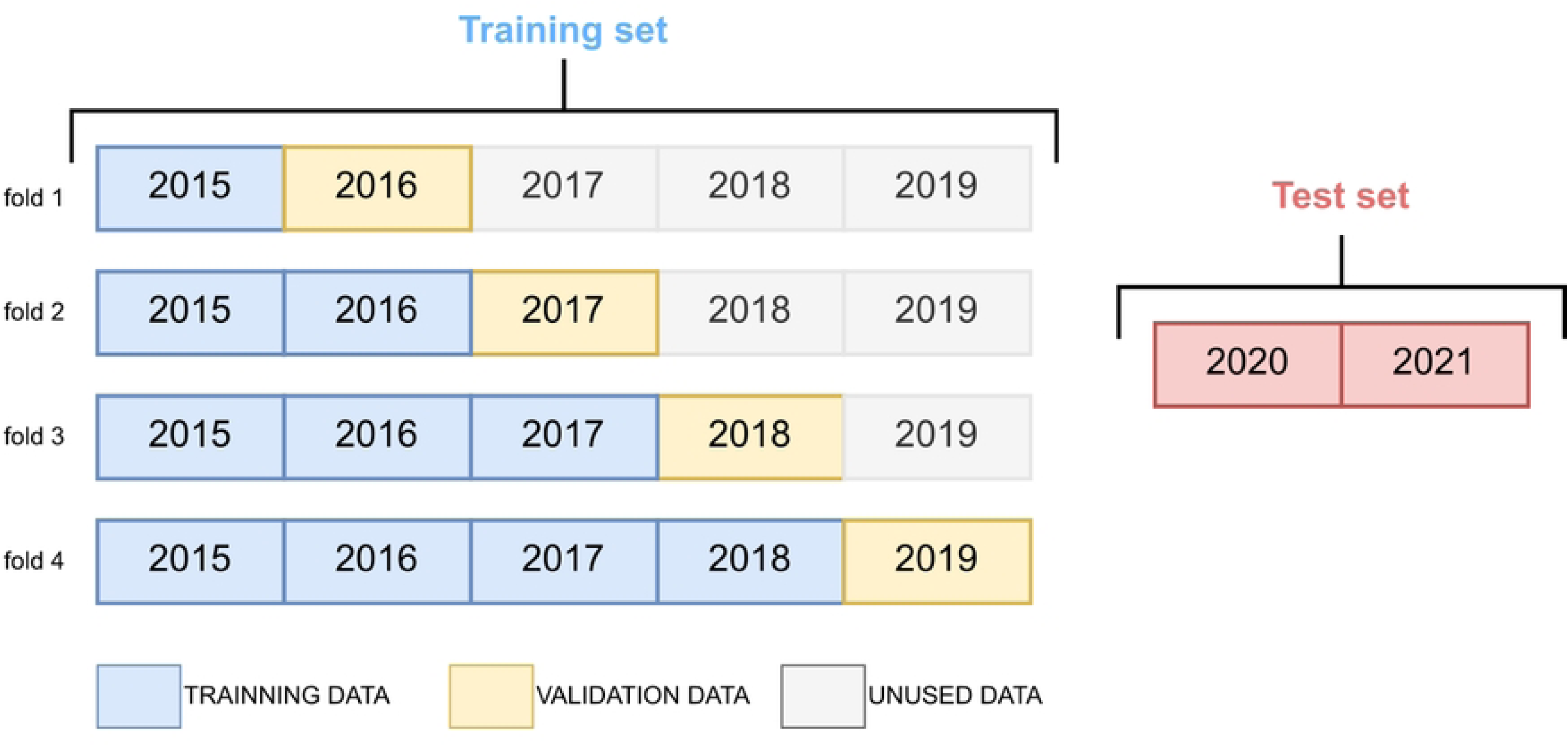
Time series split for k-fold cross validation: training and validation set.

The final selected model was a quasi-Poisson regression model that relates the logarithm of the monthly number of deaths to various time-dependent predictors, including annual trends, long-term trends, climatic variables such as average monthly maximum and minimum temperatures, monthly reports of Influenza-like Illness cases, and the month of the year. The models were fitted to the data using the GAM function in R statistical software’s *mgcv* library^31^. A separate model was fitted for each geographical area and one at the national level. A second-order autoregressive correlation structure was specified in the model to account for temporal correlation in the data. This allows the modeling of the dependence of errors over time. An examination of the residuals of the models showed a good fit to the data, including all lag autocorrelations. The annual trend was modeled using a cyclic B-spline with predefined smoothing levels.

Excess mortality percentage indicators were calculated using the selected GAM. As elaborated in this section, multivariate models and their corresponding prediction intervals were developed for each geographic area or jurisdiction to estimate expected deaths on a monthly basis for the 2020-2021 period. The excess percentages for 2020 and 2021 were calculated using the thresholds generated by this model, involving the summation of the monthly estimates for each year. Unlike previous methods, which were based solely on the mean and median of the past five years, this approach enables distinct estimations for different years. Additionally, a prediction interval was constructed from which the confidence intervals for the excess indicator were derived. These intervals were calculated using the model’s statistical estimates and the properties of the residual distribution with the predict() function from *mgcv* library.

The number of excess deaths was calculated on a monthly basis as the difference between the total observed deaths from all causes and the expected deaths based on the model. The confidence interval for the number of excess deaths was determined by using the prediction interval of the fitted model. Annual summary indicators were constructed by summing the monthly values. The p-score, or percentage of excess mortality, was calculated as the ratio of excess deaths to expected deaths per 100 per month, year, and geographic region.

Values for the entire country were computed using a stratified approach as described by Nielsen et al.^32^. Unlike the summarized approach, this approach assumes statistical independence among geographic regions and has the advantage of mitigating discrepancies between the national and regional levels.

During the development of our study, an interactive visualization was created using the R programming language, utilizing the Shiny package^33^ for the user interface and the Highcharter package^34^ for graphical representations..

The codes used to fit the models are publicly available and are accessible in the GitHub repository (https://github.com/agsantoro/excesoMortalidad). The results of this study can be visualized at https://iecs.shinyapps.io/excesomortalidad.

## Results

Figs 3, 4, and 5 present a comprehensive graphical representation of the data utilized as covariates in our models, described by province/region. Figure 3 displays the temperature variables. Figure 4 depicts the number of reported cases of Influenza-Like Illness, providing insights into the epidemiological trends across different provinces. Figure 5 charts the mortality data from all causes. These figures collectively facilitate a nuanced understanding of the interplay between environmental factors, disease incidence, and mortality outcomes.

**Fig 3.**
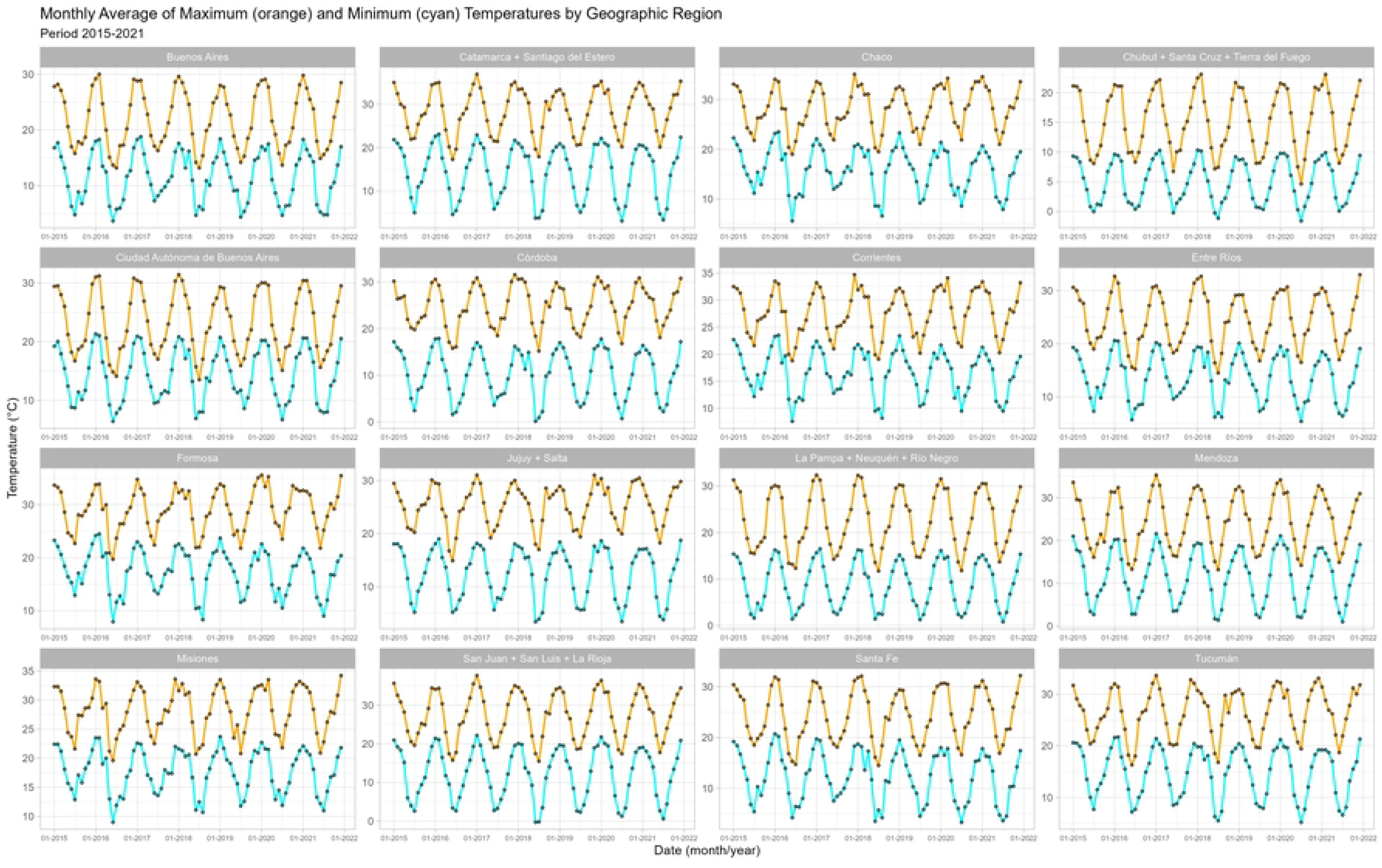
Monthly Average of Maximum (orange) and Minimum (cyan) Temperatures by Geographic Region. Period 2015-2021.

**Fig 4.**
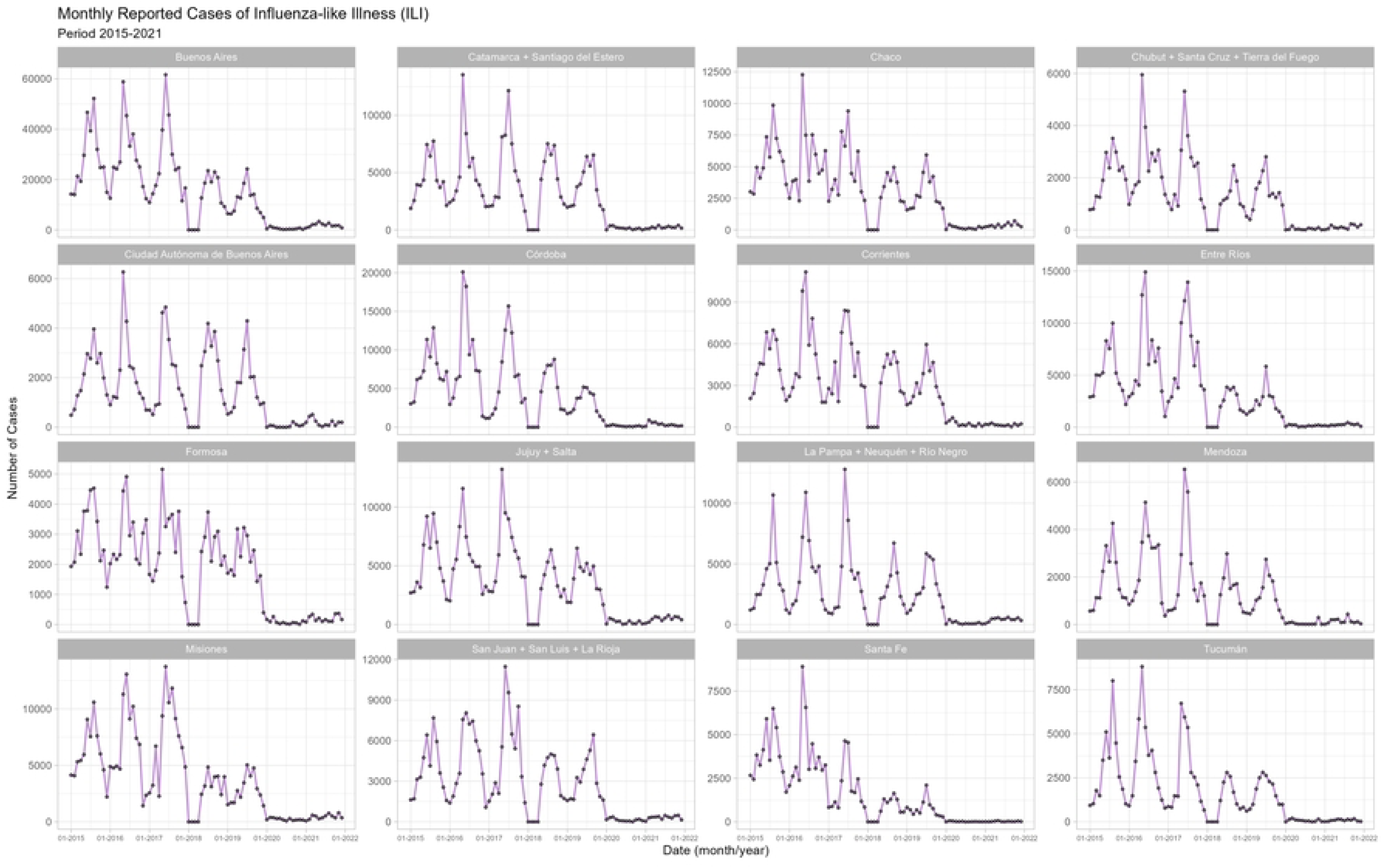
Monthly Reported Cases of Influenza-like Illness (ILI). Period 2015-2021.

**Fig 5.**
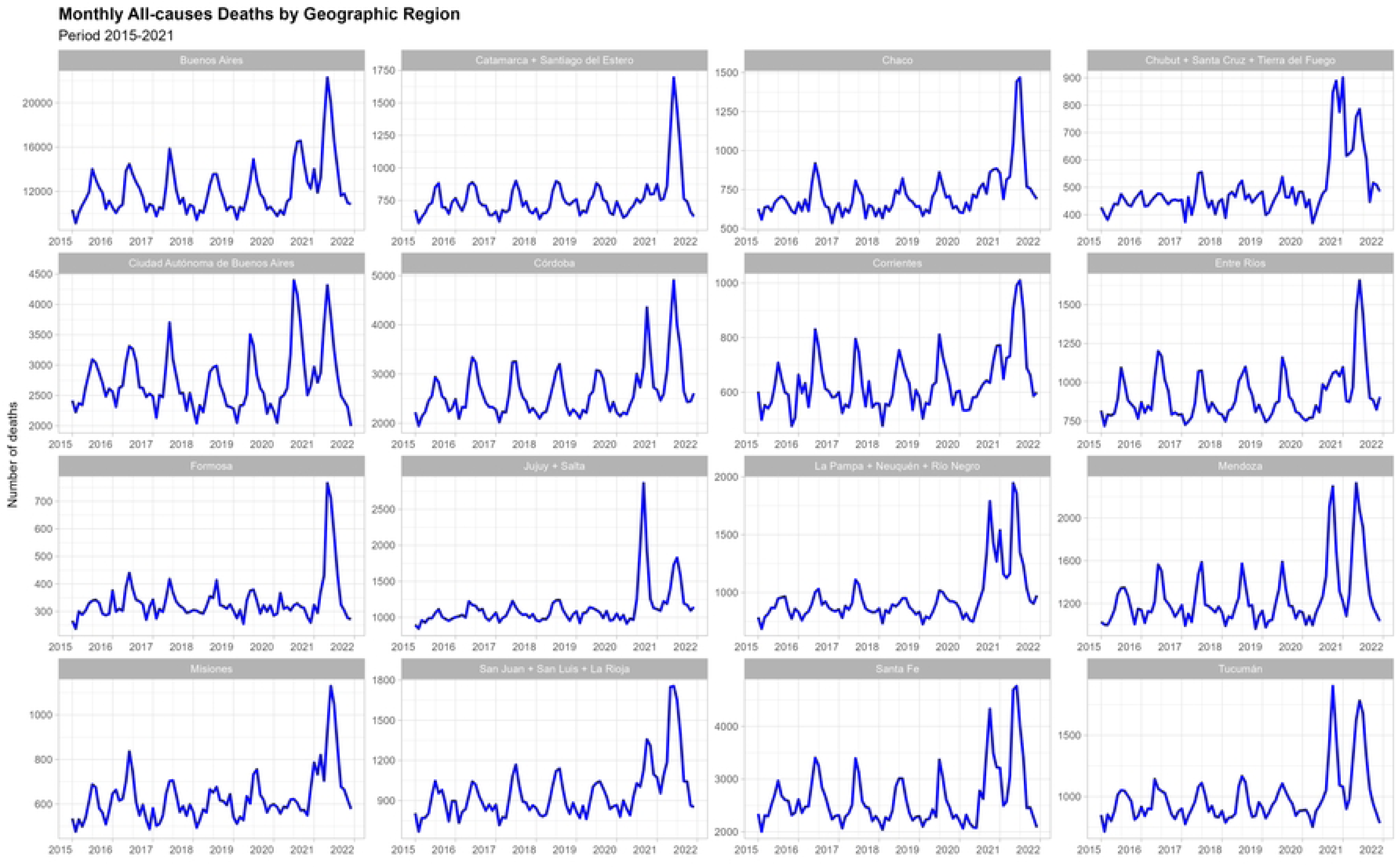
Monthly All-causes Deaths by Geographic Region. Period 2015-2021.

Based on the method employed for the entire period, 133,612 excess deaths were identified in Argentina, while 137,736 deaths are reported due to COVID-19 for the full period (2020-2021). This discrepancy yields an undercount ratio of 0.97. The undercount ratio highlights the percentage by which reported disease-specific deaths deviate from excess deaths estimated from all-cause mortality data, which in this instance suggests a 3% underreporting of COVID-19 deaths. Upon annualized assessment for the year 2021, 91,125 excess deaths are described alongside 84,190 COVID-19 deaths, resulting in a higher undercount ratio of 1.08.

### Waves identified

When observing by geographical area, it is noteworthy that in the province of Buenos Aires and the City of Buenos Aires (CABA), three peaks of deaths above expected levels were identified, with elevated mortality levels between each one. These events occurred in July, August, and September 2020; January 2021; and May 2021. Given that the province of Buenos Aires is the largest and most populated jurisdiction in the country, the same pattern of three peaks of excess deaths is found when analyzing the total for Argentina. However, in the jurisdictions of Córdoba, Entre Ríos, Santa Fe, Corrientes, Chaco, Tucumán, Mendoza, and the NOA 1, Resto Cuyo, and Patagonia Norte regions, two peaks were identified, one in 2020 and another in 2021 (months from May to June). In these geographic areas, the level of mortality did not return to the expected value during any period of the study (figure 6).

**Fig 6.**
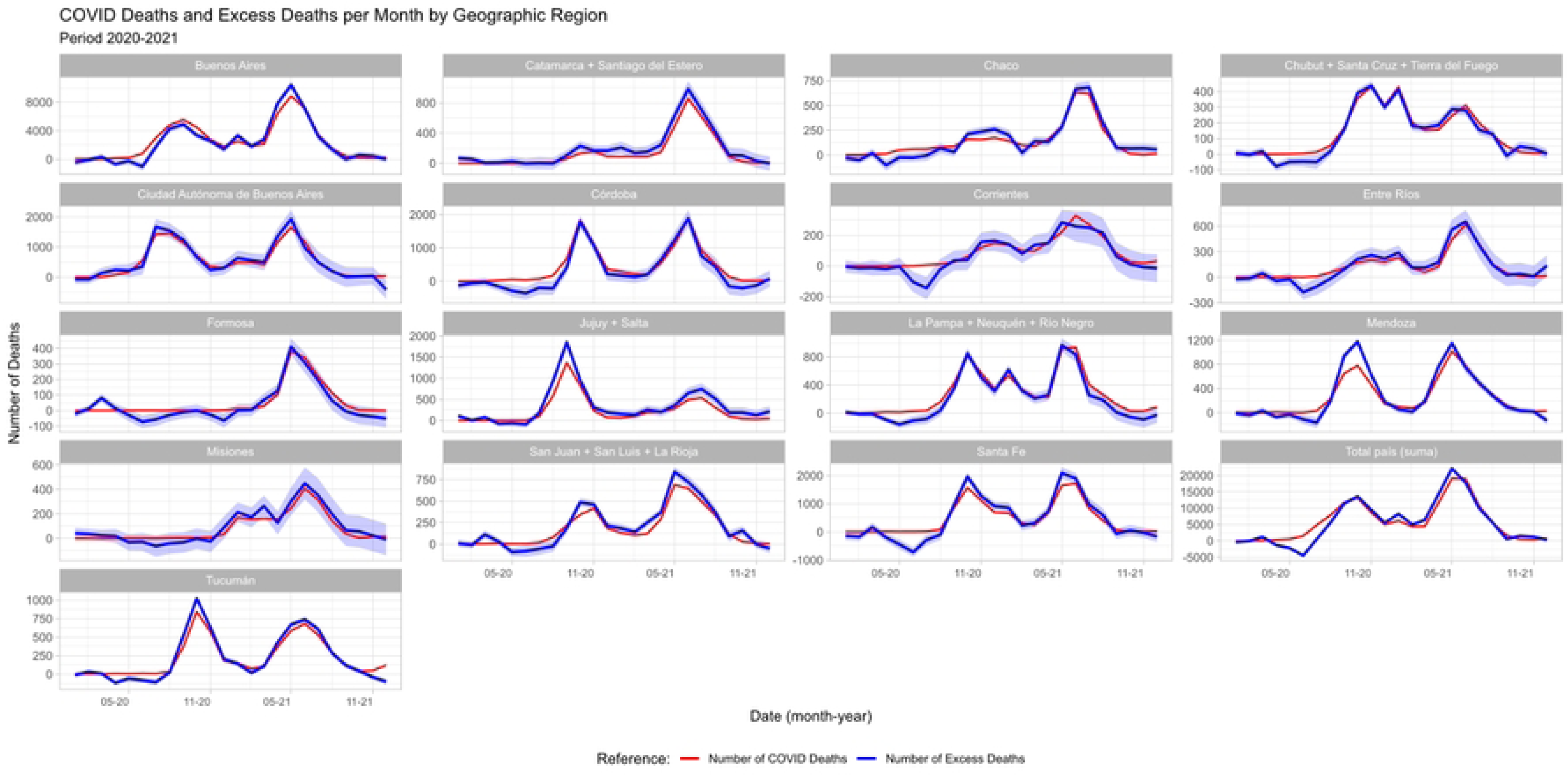
COVID Deaths and Excess Deaths per Month by Geographic Region. Period 2020-2021.

### First Half of 2020

During the first half of 2020, four geographical areas were identified with mortality levels below expected levels (Santa Fe, Córdoba, Tucumán, and Patagonia Norte, see Table 1). The rest of the areas studied did not show significant excess mortality, as the percentage excess presented a confidence interval that included zero.

**Table 1.**
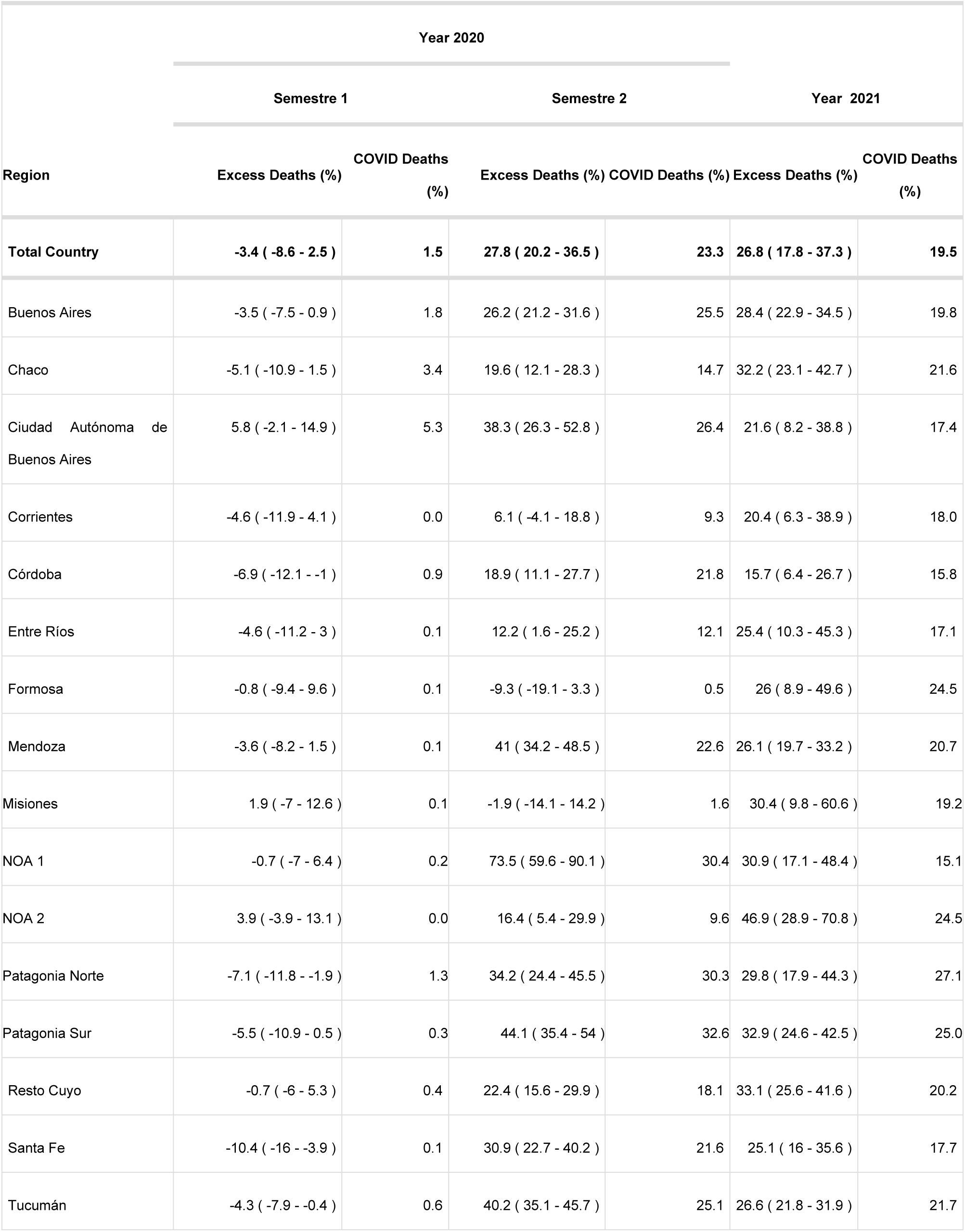

### Second Half of 2020

During the second half of 2020, following the multivariate modeling methodology, three geographical areas were identified with no significant excess mortality (Misiones, Corrientes, and Formosa). The rest of the areas studied showed significant excess mortality, with the three most affected being Noa 1 (73.5% CI 59.6 - 90.1), Patagonia Sur (44.1% CI 35.4 - 54), and Mendoza (41% CI 34.2 - 48.5). The regions with the lowest levels of excess for the second half of 2020 were Entre Rios (12.2% CI 1.6 - 25.2), NOA 2 (16.4% CI 5.4 - 29.9), and Córdoba (18.9% CI 11.1 - 27.7).

In comparing the indicator of the percentage of COVID-19 related deaths to the percentage of excess mortality, it was observed that in all the studied regions exhibiting significant excess, the excess mortality surpassed the percentage of deaths attributed to COVID-19. However, this trend was not observed in the provinces of Córdoba and Corrientes (fig 6)

### Year 2021

The year 2021 showed significant levels of excess mortality in all studied geographical areas. The highest levels of the p-score indicator were observed in the NOA 2 area, which reached values of 46.9% (CI 28.9 - 70.8), Resto de Cuyo (33.1% CI 25.6 - 41.6), Patagonia Sur (32.9%, CI 24.6 - 42.5), and Chaco (32.2, CI 23.1 - 42.7). Fig 6 illustrates the magnitude of the excess in these geographical areas. It can also be observed that, unlike 2020, excess mortality in these areas was distributed between January and September, in contrast to 2020, where it was mainly concentrated between August and December. The regions of Córdoba (15.7% (CI 6.4 - 26.7), Corrientes (20.4% (CI 6.3 - 38.9), and CABA (21.6%; CI 8.2 - 38.8) showed lower levels of excess mortality. In 2021, all geographical regions displayed higher mortality rates than those presented by COVID-19 deaths.

The interactive visualization https://iecs.shinyapps.io/excesomortalidad) displaying the results was developed to enable users from various levels and jurisdictions to conduct their own analyses, select indicators, and easily obtain results. It allows for comparisons between regions and the disaggregation of indicators at different levels. Although the models underlying the visualization are sophisticated, the platform facilitates straightforward interpretation and can be customized to meet the needs of specific users.

### Limitations

It is important to note that the excess mortality measures employed in this study (both the quantity and percentage of excess deaths) do not account for the differences in the demographic structures of the compared populations. While there are precedents for studies that have used age-adjusted indicators to estimate excess mortality, most of the studies did not incorporate this adjustment. In future approaches, it is crucial to evaluate whether to prioritize demographic structure as a determinant of the magnitude of excess (older populations tend to exhibit a higher proportion of excess deaths due to the overrepresentation of older age groups) or regarding this structure as a bias, prioritizing interpretations based on the comparative impact of the anomalous phenomenon. Additionally, it is worth noting that the excess mortality measures used in this study are not based on differences between age-adjusted mortality rates and therefore possess inherent interpretability.

The lack of mortality data for 2022 at the time this study was conducted is another limitation. In Argentina, the temporal criteria for the statistical recording of deaths are based on the date of registration; therefore, the release of 2022 data will also include deaths that occurred in 2021 (primarily towards the end of the year) but were reported to the MoH in 2022. This circumstance is evident in Fig 6, where a noticeable decrease in excess mortality is observed in November and December 2021. It should also be considered that the inclusion of these deaths will elevate the annual figures for excess deaths and excess mortality for 2021, particularly in the final months of the year, as the vast majority of deaths registered in the subsequent year, originate in November and December of the preceding year.

We should take into account some idiosyncrasies of the data used. Deaths have been tabulated by their "underlying cause of death"^35^ which was selected for each death based on rules among the multiple causes of death indicated in the death certificate. However, due to the novelty of the COVID-19 event, the WHO established new rules for its registry in 2020^36^, prioritizing COVID-19 over other causes, which could constitute a bias of over-representation of mortality due to COVID-19 and, consequently, an under-representation of mortality from other causes. In other words, deaths with COVID-19 rather than deaths by COVID-19.

## Discussion

First, based on the methodology employed in this study for the entire period, we identified 133,612 excess deaths in Argentina, while 137,736 COVID-19 deaths were reported for the entire period (2020-2021). This discrepancy yields an undercount ratio of 0.97. However, when examining each year individually, 2021 shows 91,125 excess deaths and 84,190 reported COVID-19 deaths, leading to an undercount ratio of 1.08. Argentina’s estimated undercount ratio for the entire period of 2020-2021 aligns closely with those of Chile (0.99) and Panama (1.01) for 2020, and is lower than those of Brazil (1.11), Paraguay (1.15), and Peru (1.07) for the same period^7^.

The GBD 2021 provides vital demographic estimates spanning 204 countries, including Argentina, emphasizing pandemic-period changes in mortality and life expectancy, underscoring the need for timely data to grasp COVID-19’s impact on population health trends.^3^

Notably, the excess deaths estimated for Argentina in this study are also in line with figures from other sources. The Institute for Health Metrics and Evaluation (IHME)^37^ estimates 141,488 excess deaths, while The Economist^38^ reports a figure of 132,470 for the entire period.

The challenges associated with analyzing excess mortality in Argentina are significant due to the absence of a digital death registry^8^, which impedes real-time pandemic response and delays the provision of vital statistics. Studies by Rearte et al. (2020)^8^ reveal a 10.6% increase in excess mortality in 2020 compared with the 12.7% obtained in this study, although the methodology may oversimplify outcomes by not accounting for environmental, and long -term trend and annual trends. Regional insights provided by Sarrouf et al. (2020)^10^ and Pesci et al. (2021)^39^ highlight disparate impacts across the country, with notable increases in areas like Patagonia and Buenos Aires province, reflecting healthcare saturation and altered public behaviors. Additionally, a reported decline in non-COVID-19 mortality among the elderly suggests significant behavioral and healthcare engagement changes during the pandemic.

Argentina’s struggle to manage mortality data is not unique, as a significant digital divide hinders timely public health interventions globally. The EuroMOMO^40^ initiative serves as a model for Argentina to enhance its pandemic response and data analysis capabilities. Comparative analyses by Rossen et al. (2021)^18^ highlight universal challenges and the sustained impacts of the pandemic, emphasizing the global scale of the crisis and the common hurdles in pandemic management and response strategies.

Global insights from the WHO (2020)^2^ estimate 14.8 million excess deaths worldwide, underscoring the critical need for robust data collection and analysis methodologies. The parallel between global figures and national findings in Argentina highlights the underreporting and challenges in accurately assessing the pandemic’s full impact. By aligning Argentina’s research approaches with global standards and insights, there is an opportunity to refine public health strategies, enhance real-time monitoring, and ensure a more informed and effective response to current and future public health crises.

During the COVID-19 pandemic, a significant substitution of causes of death was noted, where declines in mortality from non-COVID-19 causes were observed, with diabetes being the notable exception^9^. This phenomenon highlights the complex interplay between the pandemic and other health conditions and deserves a specific approach. In particular, the study showcases that during 2020, there was a notable decrease in mortality across various non-COVID-19 categories, illustrating the shift in mortality causes during the pandemic period. However, these changes in the mortality profile must be evaluated taking into account the particularities of each group of causes of death and the historical period (circulation restriction measures, difficulties in accessing health services, etc.).

Moreover, the investigation reveals that in 2021, excess deaths estimated by the study exceeded the number of deaths officially recorded as due to COVID-19, suggesting possible underreporting and an increase in deaths from other causes, potentially linked to the pandemic’s indirect impacts (REF). This situation aligns with the concept of syndemics, emphasizing the exacerbated mutual impacts of COVID-19 and non-communicable diseases, affecting individuals with chronic conditions. Understanding this dynamic requires a comprehensive approach, integrating the substitution phenomenon observed in 2020, where a reduction in non-COVID deaths, aside from diabetes, coincides with the pandemic’s wider implications. This context underscores the need for precise methodologies for estimating all-cause excess mortality and understanding non-COVID mortality behaviors, particularly regarding NCDs, to inform public health decisions and address the intertwined challenges of infectious and chronic diseases.

## Conclusions

To conclude, our examination of excess mortality in Argentina during the COVID-19 pandemic provides significant insights into the extensive impact of the pandemic, beyond just the reported infection and mortality rates. These insights greatly contribute to our understanding of the pandemic’s broader effects and emphasize the critical need for detailed public health responses. Highlighting how pandemic preparedness and response strategies can be informed by the lessons learned from the COVID-19 mortality rates, we see a clear directive for strengthening health systems and enhancing response mechanisms to better manage future public health crises. Additionally, it is crucial to extend our research by analyzing excess mortality due to different causes of death. Further analysis will improve our comprehension of the pandemic’s complex effects and help in crafting focused health interventions. Future studies should track these developments using thorough methods to support the ongoing and future public health efforts.

## Data Availability

All data underlying the findings of our study are available without restriction. The datasets generated and analyzed during our research are hosted on GitHub. This includes raw data, processed data, and the code used for analysis in the study titled “Assessing Excess Mortality Patterns in Argentina over the COVID-19 Pandemic (2020-2021): A Comprehensive National and Subnational Analysis.” The data and resources can be accessed at the following URL: https://github.com/agsantoro/excesoMortalidad. There are no ethical or legal restrictions on data sharing for this study. All data are fully anonymized and do not include any information that could compromise the confidentiality or privacy of the individuals represented in the dataset.

## Notes

### Competing Interest Statement

The authors have declared no competing interest.

### Funding Statement

This research was completely supported by the Centro de Implementación e Innovación en Políticas de Salud (CIIPS) at the Instituto de Efectividad Clínica y Sanitaria (IECS), Buenos Aires, Argentina. While CIIPS provided institutional support, no specific grant numbers are applicable as there was no direct external funding allocated specifically for this study. No commercial companies funded the study or the authors, and no salaries or other funding were received from commercial entities in relation to this work. As authors based in a low and middle-income country (LMIC), we would like to request a partial waiver for the publication fees. All aspects of the research were conducted independently by the authors.

## References

1. WHO Coronavirus (COVID-19) Dashboard. https://covid19.who.int/.

2. Msemburi, W. et al. The WHO estimates of excess mortality associated with the COVID-19 pandemic. Nature 613, 130–137 (2023).

3. Schumacher, A. E. et al. Global age-sex-specific mortality, life expectancy, and population estimates in 204 countries and territories and 811 subnational locations, 1950--2021, and the impact of the COVID-19 pandemic: a comprehensive demographic analysis for the Global Burden of Disease Study 2021. Lancet (2024).

4. Reporte interactivo de estadísticas de salud. Argentina.gob.ar https://www.argentina.gob.ar/salud/deis/reporte-interactivo (2021).

5. Gideon, J. Introduction to COVID -19 in Latin America and the Caribbean. Bull. Lat. Am. Res. 39, 4–6 (2020).

6. Karlinsky, A. & Kobak, D. The World Mortality Dataset: Tracking excess mortality across countries during the COVID-19 pandemic. medRxiv (2021) doi:10.1101/2021.01.27.21250604.

7. Lima, E. E. C. et al. Investigating regional excess mortality during 2020 COVID-19 pandemic in selected Latin American countries. Genus 77, 30 (2021).

8. Pennini, V. & Santoro, A. Exceso de mortalidad durante la pandemia de COVID-19 en Argentina. GitHub/excesoMortalidad https://github.com/agsantoro/excesoMortalidad.

9. Sarrouf, E. B., Marconi, A. & Cuezzo, M. R. Modificaciones en la mortalidad por causas diferentes a COVID-19 durante el primer año de pandemia en Argentina, 2020. Rev Argent Salud Pública 15, e108–e108 (2023).

10. Sarrouf, E. B. Exceso de mortalidad por todas las causas en la Argentina y sus 24 jurisdicciones, 2020. REVISTA ARGENTINA DE MEDICINA 10, 174–174 (2022).

11. Knutson, V., Aleshin-Guendel, S., Karlinsky, A., Msemburi, W. & Wakefield, J. Estimating global and country-specific excess mortality during the Covid-19 pandemic. aoas 17, 1353–1374 (2023).

12. Palacio Mejía, L. S., et al. [Not Available]. Salud Publica Mex. 63, 211–224 (2021).

13. Vieira, A., Peixoto, V. R., Aguiar, P. & Abrantes, A. Rapid Estimation of Excess Mortality during the COVID-19 Pandemic in Portugal -Beyond Reported Deaths. J. Epidemiol. Glob. Health 10, 209–213 (2020).

14. Blangiardo, M. et al. Estimating weekly excess mortality at sub-national level in Italy during the COVID-19 pandemic. PLoS One 15, e0240286 (2020).

15. Scortichini, M. et al. Excess mortality during the COVID-19 outbreak in Italy: a two-stage interrupted time-series analysis. Int. J. Epidemiol. 49, 1909–1917 (2021).

16. COVID-19 Excess Mortality Collaborators. Estimating excess mortality due to the COVID-19 pandemic: a systematic analysis of COVID-19-related mortality, 2020-21. Lancet 399, 1513–1536 (2022).

17. Todd, M., Pharis, M., Gulino, S. P., Robbins, J. M. & Bettigole, C. Excess Mortality During the COVID-19 Pandemic in Philadelphia. Am. J. Public Health 111, 1352–1357 (2021).

18. Rossen, L. M. et al. Excess all-cause mortality in the USA and Europe during the COVID-19 pandemic, 2020 and 2021. Sci. Rep. 12, 18559 (2022).

19. Peretz, C. et al. Excess mortality in Israel associated with COVID-19 in 2020-2021 by age group and with estimates based on daily mortality patterns in 2000-2019. Int. J. Epidemiol. 51, 727–736 (2022).

20. Alfaro, T., Martinez-Folgar, K., Vives, A. & Bilal, U. Excess Mortality during the COVID-19 Pandemic in Cities of Chile: Magnitude, Inequalities, and Urban Determinants. J. Urban Health 99, 922–935 (2022).

21. de la Nación, M. de S. Defunciones Generales Mensuales ocurridas y registradas en la República Argentina. Cantidad de defunciones Generales Mensuales ocurridas y registradas en la República Argentina. Incluye Variables Sociodemográficas: Región, jurisdicción, año y mes de defunción, mes de defunción, año de defunción, sexo, grupo etario, causa de defunción, y cantidad.

22. Gasparrini, A. et al. Mortality risk attributable to high and low ambient temperature: a multicountry observational study. Lancet 386, 369–375 (2015).

23. Barnett, A. G., Tong, S. & Clements, A. C. A. What measure of temperature is the best predictor of mortality? Environ. Res. 110, 604–611 (2010).

24. Servicio Meteorologico Nacional. https://www.smn.gob.ar/.

25. Thompson, W. W. et al. Estimates of US influenza-associated deaths made using four different methods. Influenza Other Respi. Viruses 3, 37–49 (2009).

26. Moriyama, M., Hugentobler, W. J. & Iwasaki, A. Seasonality of Respiratory Viral Infections. Annu Rev Virol 7, 83–101 (2020).

27. de la Nación, M. de S. Boletines epidemiológicos. argentina.gob.ar https://bancos.salud.gob.ar/bancos/materiales-para-equipos-de-salud/soporte/boletines-epidemiologicos/.

28. de la Nación, M. de S. Vigilancia de Infecciones respiratorias agudas. Datos Abiertos del Ministerio de Salud http://datos.salud.gob.ar/dataset/vigilancia-de-infecciones-respiratorias-agudas.

29. Foundation for Statistical Computing, R. R. R: A language and environment for statistical computing. RA Lang Environ Stat Comput.

30. Kuhn & Max. Building Predictive Models in R Using the caret Package. Journal of Statistical Software vol. 28 1–26 Preprint at 10.18637/jss.v028.i05 (2008).

31. Wood, S. N. Generalized Additive Models: An Introduction with R, Second Edition. (CRC Press, 2017).

32. Nielsen, J. et al. Pooling European all-cause mortality: methodology and findings for the seasons 2008/2009 to 2010/2011. Epidemiol. Infect. 141, 1996–2010 (2013).

33. Chang, W., et al. shiny: Web Application Framework for R. Preprint at https://CRAN.R-project.org/package=shiny (2023).

34. Kunst, J. highcharter: A Wrapper for the ‘Highcharts’ Library. Preprint at https://CRAN.R-project.org/package=highcharter (2022).

35. Cause of death. https://www.who.int/standards/classifications/classification-of-diseases/cause-of-death.

36. Singh, B. International comparisons of COVID-19 deaths in the presence of comorbidities require uniform mortality coding guidelines. Int. J. Epidemiol. 50, 373–377 (2021).

37. Homepage. Institute for Health Metrics and Evaluation https://www.healthdata.org/.

38. The. Tracking covid-19 excess deaths across countries. The Economist (2021).

39. Pesci, S. et al. Exceso de mortalidad por la pandemia de COVID-19 durante 2020 en la provincia de Buenos Aires, Argentina. Rev. Arg. Salud Pública 13, (2021).

40. Mølbak, K. & Mazick, A. European monitoring of excess mortality for public health action (EuroMOMO) Kåre Mølbak. Eur. J. Public Health 23, ckt126–113 (2013).

